# Development and Validation of the TikTok Addiction Scale-Short Form (TTAS-SF)

**DOI:** 10.1101/2025.11.24.25340870

**Authors:** P. Galanis, A. Katsiroumpa, O. Konstantakopoulou, O. Galani, M. Tsiachri, I. Moisoglou

**Author notes:** Corresponding author: P. Galanis, 123 Papadiamantopoulou street, GR-11527, Athens, Greece. **Funding** None. **Competing interests** None. **Ethical approval** All procedures followed were in accordance with the ethical standards of the responsible committee on human experimentation (institutional and national) and with the Helsinki Declaration of 1975, as revised in 2000. Informed consent was obtained from all participants for being included in the study.

## Abstract

**OBJECTIVE:** To develop and validate a short version of the TikTok Addiction Scale, i.e. the TikTok Addiction Scale-Short Form (TTAS-SF).

**METHOD:** Construct validity of the TTAS-SF was assessed through corrected item–total correlations and confirmatory factor analysis. Concurrent validity was examined using the Bergen Social Media Addiction Scale (BSMAS), the Patient Health Questionnaire-4 (PHQ-4), and the Big Five Inventory-10 (BFI-10). Reliability was evaluated through multiple indices, including Cronbach’s alpha, McDonald’s omega, Cohen’s kappa, and the intraclass correlation coefficient.

**RESULTS:** Corrected item–total correlations and confirmatory factor analysis confirmed that the final version of the TTAS-SF includes six items in one factor. Concurrent validity of the TTAS-SF was excellent since we found statistically significant correlations between the TTAS-SF and the BSMAS, the PHQ-4, and the BFI□10. Cronbach’s alpha and McDonald’s Omega for the TTAS-SF was 0.805 and 0.817, respectively. Cohen’s kappa for the six items ranged from 0.789 to 0.905 (p < 0.001 in all cases). Additionally, intraclass correlation coefficient was 0.993 (p < 0.001). Thus, the reliability of the TTAS-SF was excellent. The best cut-off point for the TTAS-SF was 13, indicating that TikTok users with TTAS-SF score ≥13 were considered as users with a problematic TikTok use and high probability of addiction issues, and those with TTAS-SF score <13 as healthy users.

**CONCLUSIONS:** The TTAS-SF is a one-factor 6-item scale with great reliability and validity. The TTAS-SF is a short and easy-to-use tool that measures levels of TikTok addiction in a couple of minutes. Valid measurement of TikTok addiction with brief and valid tools is essential to further understand predictors and consequences of this phenomenon.

## Introduction

TikTok has emerged as one of the most widely used platforms for short-form video content globally. Approximately one in five individuals aged 18 years and older use TikTok, and the application was downloaded over two billion times in 2021. The majority of its users are adolescents and young adults, typically between 16 and 35 years of age. While TikTok offers a free platform for creating, editing, and sharing short video clips enhanced with filters and trending music, concerns persist regarding the potential negative consequences of social media use. Unlike other social networking platforms such as Facebook, Twitter, Instagram, and Snapchat, which primarily emphasize images and text, TikTok is distinguished by its focus on brief video content.(1)

The excessive use of social media is becoming a significant public health issue due to its association with various problems, such as depression, low self-esteem, impulsivity, suicide risk, work impairments, and poor sleep quality.(2–7) TikTok enables users to capture memorable moments and create concise videos that document their lives, offering substantial entertainment value. However, it has also emerged as a potential source of social media addiction. Social media addiction is characterized by the repeated occurrence of addiction-like symptoms and a diminished capacity for self-control in relation to social media use.(8) Social media addiction is defined as the persistent experience of addiction-like symptoms or a diminished ability to exercise self-control in relation to social media use.(9,10)

Recent evidence suggests the negative impact of problematic TikTok use on several issues, such as anxiety, depression, loneliness, poor sleep, procrastination and self-esteem.(11–14) Although existing research on social media addiction has primarily focused on platforms such as Facebook, Instagram, and other well-established networks, the influence of TikTok and its associated maladaptive behaviors has received comparatively little attention.(8)

Although numerous instruments exist to assess social media, social networking, internet, Facebook, and Instagram addiction or problematic use,(15) there is a notable lack of valid and specific psychometric tools designed to evaluate TikTok addiction. A recent scoping review found that there are 37 instruments that measure negative social networking site The Bergen Facebook Addiction Scale (BFAS) is the most widely instrument for measuring negative use of social media.(16)

Given the rapid growth in TikTok usage and the possibility that TikTok addiction represents a distinct form of social media addiction, the development of a reliable measurement tool is imperative. Furthermore, considering the diversity in design and functionality across social media platforms, it is essential to investigate the impact of TikTok use on individuals’ mental health. Different platforms may exert varying influences on users, potentially resulting in unique outcomes for psychological well-being.

Thus, recently, the TikTok Addiction Scale (TTAS)(17) is developed to measure in a reliable and valid way levels of TikTok addiction. Given that the TTAS comprises 15 items, the development of a shorter, yet valid, version of the scale could provide a more efficient and accessible tool for measuring TikTok addiction. Thus, the aim of this study was to develop and validate a short version of the TikTok Addiction Scale, i.e. the TikTok Addiction Scale-Short Form (TTAS-SF).

## Methods

### Study design

A cross-sectional study was conducted using an online survey, with data collected in July 2024. The inclusion criteria were as follows: (a) participants had to be active TikTok users for at least the preceding 12 months, (b) participants were required to be adults, and (c) informed consent had to be obtained prior to participation.

We collected our data in an anonymous and voluntary basis. We informed participants about the aim and the design of our study and they gave their informed consent. The Ethics Committee of the Faculty of Nursing, National and Kapodistrian University of Athens approved our study protocol (approval number; 451, June 2023). Moreover, we conducted our study in accordance with the Declaration of Helsinki.(18)

### Construct validity

The initial version of the TTAS includes 15 items and six factors; salience (two items), mood modification (two items), tolerance (three items), withdrawal symptoms (two items), conflict (four items), and relapse (two items).(17)

In this study, we considered a one-factor structure for the TTAS to obtain the TikTok Addiction Scale-Short Form. Thus, we retained in the TTAS-SF the item from each factor with the highest corrected item-total correlation. In this way, we kept the six core components of addiction, i.e., salience, mood modification, tolerance, withdrawal symptoms, conflict and relapse, by reducing the total number of the items from 15 to 6.

We employed confirmatory factor analysis (CFA) to examine the construct validity of the TTAS-SF. We calculated the following indices of fit: chi-square/degree of freedom (x^2^/df), root mean square error of approximation (RMSEA), goodness of fit index (GFI), normed fit index (NFI), and comparative fit index (CFI). Acceptable values for fit indices are the following: RMSEA < 0.10, GFI > 0.90, NFI > 0.90, CFI > 0.90, and x^2^/df < 5.(19–22)

### Concurrent validity

Concurrent validity of the TTAS-SF was investigated using the Bergen Social Media Addiction Scale (BSMAS),(23) the Patient Health Questionnaire-4 (PHQ-4),(24) and the Big Five Inventory□10 (BFI□10)(25).

Total score on the BSMAS ranges from 6 to 30, and higher scores indicate greater social media addiction. We used the valid Greek version of the BSMAS.(26) In our study, Cronbach’s alpha for the BSMAS was 0.829, and McDonald’s Omega was 0.829.

Total score on the PHQ-4 ranges from 0 to 12, and higher scores indicate greater anxiety and depression. We used the valid Greek version of the PHQ-4.(27) In our study, Cronbach’s alpha for the PHQ-4 was 0.818, and McDonald’s Omega was 0.825.

The BFI□10 includes five factors: neuroticism, extraversion, openness, agreeableness, and conscientiousness. Total score for each factor ranges from 2 to 10. Higher scores indicate greater neuroticism, extraversion, openness, agreeableness, and conscientiousness. We used the valid Greek version of the BFI□10.(28) In our study, Cronbach’s alpha for the BFI□10 was 0.712, and McDonald’s Omega was 0.714.

We measured the total score on the TTAS-SF by adding answers on six items. Answers on the six items are on a 5-point Liker scale from 1 to 5; 1 (very rarely), 2 (rarely), 3 (sometimes), 4 (often), and 5 (very often). Thus, the total score on the TTAS-SF ranges from 5 to 30. Higher scores on the TTAS-SF indicate greater levels of TikTok addiction.

Afterwards, we expected a positive correlation between the TTAS-SF and the BSMAS, the PHQ-4, and neuroticism. Moreover, we expected a negative correlation between the TTAS-SF and extraversion, openness and conscientiousness.

### Reliability

Reliability analyses were conducted by calculating Cronbach’s alpha and McDonald’s omega for the TTAS-SF, with acceptable thresholds set at >0.60.(29) Additionally, corrected item–total correlations and Cronbach’s alpha if item deleted were computed for the 6 items of the TTAS-SF, with acceptable item–total correlation values defined as ≥0.30.(30) A test–retest reliability assessment was performed with a subsample of 30 TikTok users, who completed the TTAS-SF twice within a one-week interval. For ordinal item responses, Cohen’s kappa was calculated for the six items of the TTAS-SF. Furthermore, a two-way mixed intraclass correlation coefficient (absolute agreement) was computed for the TTAS-SF total score.

### Cut-off point

Receiver Operating Characteristic (ROC) analysis was employed to determine the optimal cut-off point for the TTAS-SF, using the Patient Health Questionnaire-4 as external criterion. Dichotomous variables for PHQ-4 were created based on cut-off points recommended in the literature: 3 for anxiety, and 3 for depression.

We calculated sensitivity, specificity, and the Youden index. These measures take values from 0 to 1 with higher values indicating better diagnostic value of the TTAS-SF. The Youden index defines an optimal cut-off point and is calculated as Sensitivity + Specificity – 1. Additionally, we calculated the area under the curve (AUC), 95% confidence interval (CI), and p-value. The best values for the Youden index and the AUC were produced by using the depression factor as the gold standard.

### Statistical analysis

Categorical variables were summarized using absolute frequencies and percentages, while continuous variables were presented as means, standard deviations, medians, and minimum and maximum values. The Kolmogorov–Smirnov test and Q–Q plots were employed to assess the distribution of scale scores. The scores for TTAS-SF, BSMAS, PHQ-4, and BFI-10 demonstrated normal distribution; therefore, Pearson’s correlation coefficient was calculated to examine the relationships among these scales. P-values less than 0.05 were considered as statistically significant. We performed CFA with AMOS version 21 (Amos Development Corporation, 2018). All other analyses were conducted with IBM SPSS 28.0 (IBM Corp. Released 2012. IBM SPSS Statistics for Windows, Version 21.0. Armonk, NY: IBM Corp.).

## Results

### Participants

The final sample consisted of 429 TikTok users. Among the participants, 81.8% were female (n = 351) and 18.2% were male (n = 78). The mean age of the sample was 26.5 years (standard deviation = 8.5), with a median age of 22 years (range: 18–54 years). Participants reported an average daily TikTok usage of 2.2 hours (standard deviation = 1.6), with a median of 2 hours and a range from 15 minutes to 8 hours.

### Construct validity

Corrected item-total correlations for the 15 items of the original version of the TTAS are shown in Table 1. We kept in the TTAS-SF the items with the highest corrected item-total correlation in each factor. Thus, for factor salience we kept the item #2, for factor mood modification, we kept the item #4, for the factor tolerance we kept the item #6, for the factor withdrawal symptoms we kept the item #9, for the factor conflict we kept the item #12, and for the factor relapse we kept the item #15.

**Table 1.** Corrected item-total correlations for the 15 items of the original version of the TikTok Addiction Scale (n=429).

| Items | Corrected item-total correlation |
| --- | --- |
| 1. I think about how I could reduce my work/study to spend more time on TikTok | 0.471 |
| **2. I have TikTok in my mind even when I am not using it** | **0.605** |
| 3. I feel calm when I use TikTok | 0.333 |
| **4. I use TikTok as a get away from my problems and my thoughts** | **0.569** |
| 5. I have had difficulties controlling the time I spend on TikTok | 0.687 |
| **6. I have had difficulties closing TikTok** | **0.701** |
| 7. I want to use TikTok more and more | 0.608 |
| 8. I feel bad when I cannot use TikTok for some time | 0.567 |
| **9. I feel sad when I cannot use TikTok for some time** | **0.571** |
| 10. I don't get enough time to do things I want to do because I spend a lot of time on TikTok | 0.667 |
| 11. I lose sleep due to excessive use of TikTok | 0.635 |
| **12. I am not able to concentrate on my work/study due to TikTok use** | **0.756** |
| 13. I use TikTok so much that it has had a negative impact on my work/study | 0.691 |
| 14. I feel depressed when I do not use TikTok, which goes when I use it | 0.583 |
| **15. I feel anxious when I do not use TikTok, which goes when I use it** | **0.619** |

Bold indicate the item with the highest corrected item-total correlation in each factor.

### Confirmatory factor analysis

Then, we performed CFA to verify the one-factor six-item model of the TTAS-SF. Our CFA suggested that the one-factor model with 6 items of the TTAS-SF had very good fit to data since x^2^/df was 2.093, RMSEA was 0.051, GFI was 0.987, NFI was 0.979, and CFI was 0.989. Standardized regression weights between the six items ranged from 0.59 to 0.73. CFA of the TTAS-SF is shown in Figure 1.

**Figure 1.**
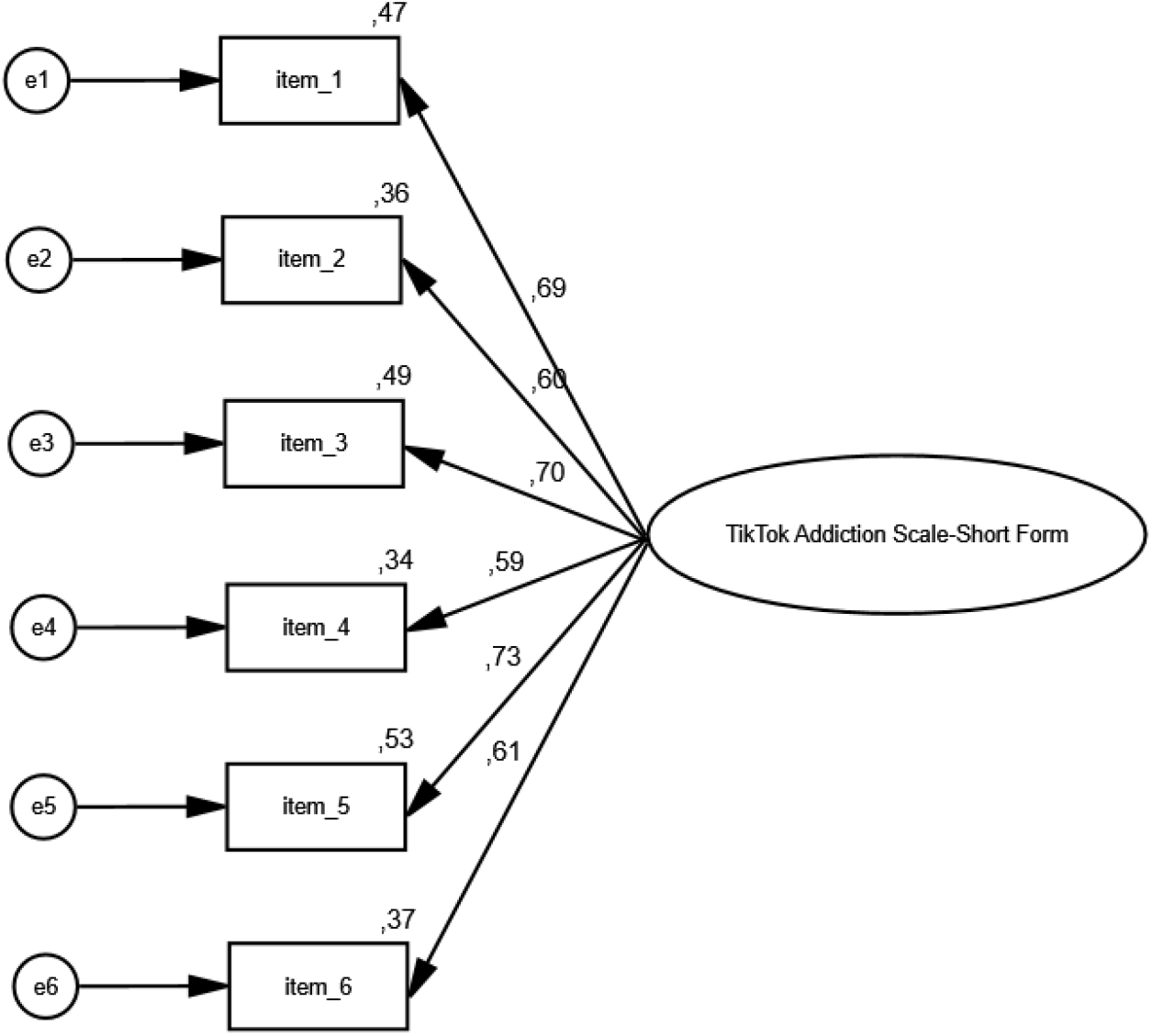
Confirmatory factor analysis of the TikTok Addiction Scale-Short Form.

Finally, the TTAS-SF is a one-factor six-item tool; salience (item #1), mood modification (items #2), tolerance (item #3), withdrawal symptoms (item #4), conflict (items #5), and relapse (item #6) (Table 2).

**Table 2.**
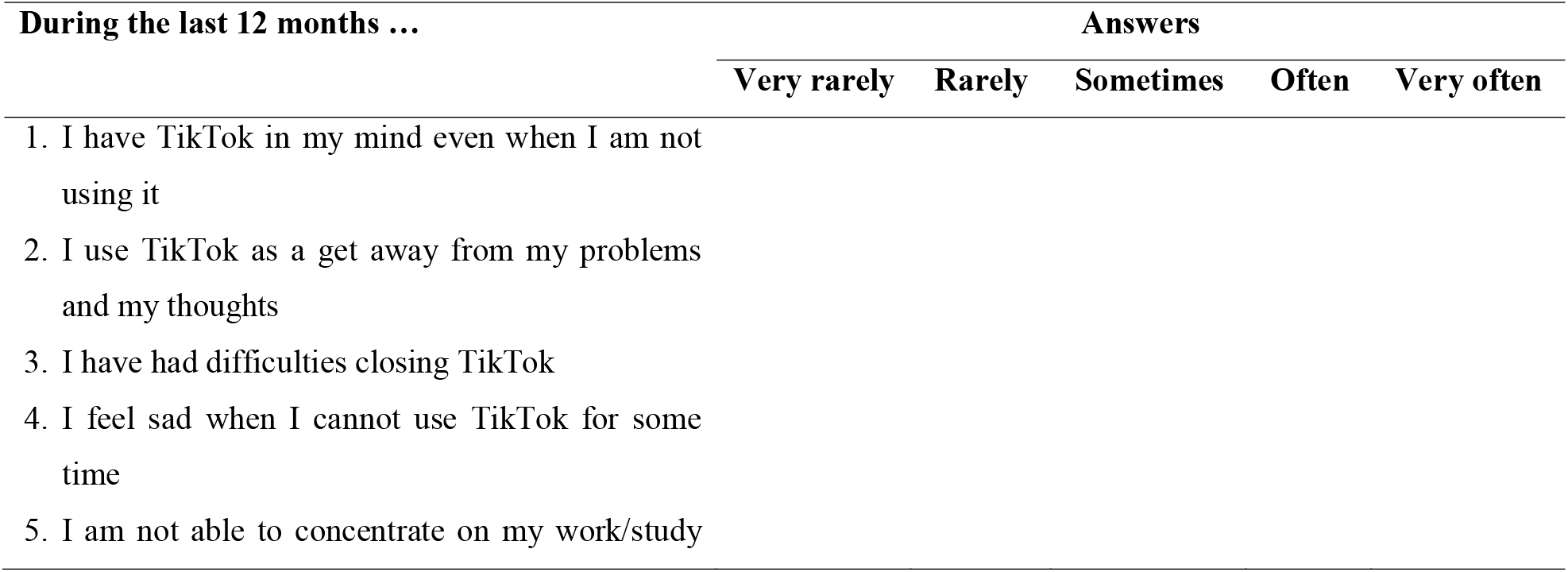
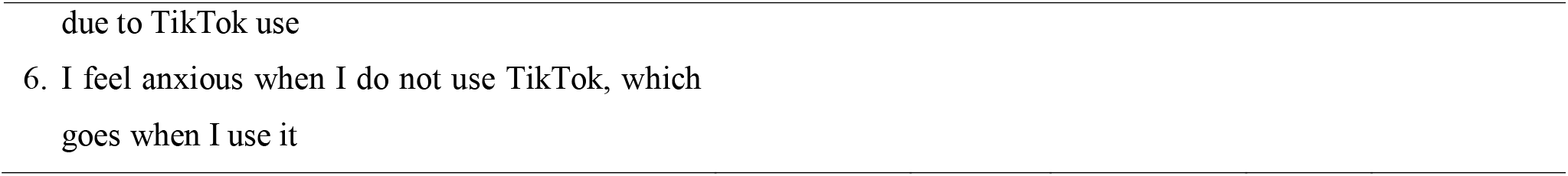
Final version of the TikTok Addiction Scale-Short Form.

### Concurrent validity

The TTAS-SF demonstrated strong concurrent validity. A positive correlation was observed between TTAS-SF and BSMAS, indicating that individuals with higher levels of social media addiction also exhibited higher levels of TikTok addiction. Similarly, TTAS-SF scores were positively correlated with PHQ-4, suggesting that participants experiencing greater symptoms of anxiety and depression tended to report higher levels of TikTok addiction. Furthermore, TTAS-SF was positively associated with neuroticism, while negative correlations were found with extraversion and conscientiousness, implying that individuals with lower levels of these traits were more likely to exhibit TikTok addiction. Table 3 presents the correlation coefficients between TTAS-SF and BSMAS, PHQ-4, and BFI-10.

**Table 3.** Pearson’s correlation coefficients between the TikTok Addiction Scale-Short Form (TTAS-SF) and the Bergen Social Media Addiction Scale (BSMAS), the Patient Health Questionnaire-4 (PHQ-4), and the Big Five Inventory□10 (BFI□10) (n=429).

| Scales | BSMAS | PHQ-4 | BFI-10 |  |  |  |
| --- | --- | --- | --- | --- | --- | --- |
|  |  |  | Neuroticism | Openness | Extraversion | Conscientiousness |
| TTAS-SF | 0.731** | 0.361** | 0.100* | -0.062 | -0.196** | -0.410** |

\* $p < 0.05$
\*\* $p < 0.01$

### Reliability

Cronbach’s alpha and McDonald’s Omega for the TTAS-SF was 0.805 and 0.817, respectively. Additionally, corrected item-total correlations had values between 0.558 and 0.647, while removal of each single item did not increase Cronbach’s alpha. Cohen’s kappa for the six items ranged from 0.789 to 0.905 (p < 0.001 in all cases). Additionally, intraclass correlation coefficient was 0.993 (95% confidence interval; 0.985 to 0.996, p < 0.001). Thus, the reliability of the TTAS-SF was excellent.

### Cut-off point

We employed ROC analysis to define an optimal cut-off point for the TTAS-SF. We found that the best cut-off point for the TTAS-SF was 13 using the factor depression of the PHQ-4 as criterion since it presented the best discriminant ability (Figure 2). In particular, we found the highest values for Youden’s index (0.378) and AUC (0.730). The value for the AUC indicated high accuracy for the cut-off point of 13. The 95% confidence interval for the AUC ranged from 0.679 to 0.781. Therefore, we considered TikTok users with TTAS-SF score ≥13 as TikTok users with a problematic TikTok use and high probability of addiction issues, and those with TTAS-SF score <13 as healthy users.

**Figure 2.**
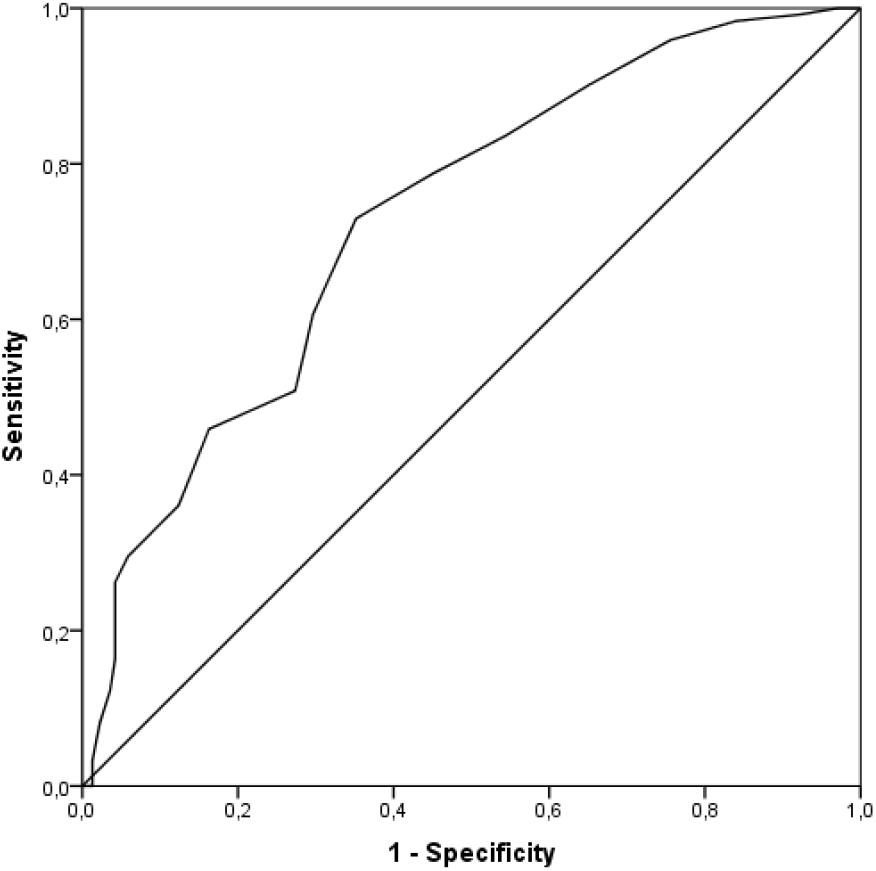
ROC curve of the TikTok Addiction Scale-Short Form.

## Discussion

Since the TikTok Addiction Scale comprises 15 items, a shorter and valid version could be an invaluable option for scholars and individuals to measure levels of TikTok addiction in just two minutes. Thus, the aim of our study was to develop and validate a short version of the TikTok Addiction Scale, i.e. the TikTok Addiction Scale-Short Form. After a thorough reliability and validity analysis, we found that the TTAS-SF is a brief and robust scale with excellent psychometric properties. After all, we found that the TTAS-SF is a one-factor 6-item scale with great reliability and validity.

Our confirmatory factor analysis verified the one-factor six-item model of the TTAS-SF. Our analysis suggested that the one-factor model with 6 items of the TTAS-SF had very good fit to data since x^2^/df was 2.093, RMSEA was 0.051, GFI was 0.987, NFI was 0.979, and CFI was 0.989.(19–22) Additionally, the correlation coefficients between the six factors ranged from 0.382 to 0.703 and were statistically significant (p<0.001 in all cases). Thus, our factor analysis identified a one-factor six-item model for the TTAS-SF, and confirmed our hypothesis that TikTok addiction involves six components (i.e., salience, mood modification, tolerance, withdrawal, conflict and relapse) as other addictions involve.

Additionally, we examined the concurrent validity of the TTAS-SF by estimating the correlation between the TTAS-SF and the BSMAS(23), the PHQ-4(24), and the BFI□10(25). The TTAS-SF demonstrated strong concurrent validity. A positive correlation was observed between TTAS-SF and BSMAS, indicating that individuals with higher levels of social media addiction also exhibited higher levels of TikTok addiction. Similarly, TTAS-SF scores were positively correlated with PHQ-4, suggesting that participants experiencing greater symptoms of anxiety and depression tended to report higher levels of TikTok addiction. Furthermore, TTAS-SF was positively associated with neuroticism, while negative correlations were found with extraversion and conscientiousness, implying that individuals with lower levels of these traits were more likely to exhibit TikTok addiction. These findings are consistent with existing literature, as two recent systematic reviews reported a positive association between excessive social media use and symptoms of depression and anxiety.(31,32) Furthermore, our results indicated that higher TTAS scores were associated with higher levels of neuroticism, aligning with previous studies that have demonstrated a positive correlation between social media addiction and neuroticism.(33–35) Additionally, individuals with elevated conscientiousness tend to prioritize work-related responsibilities over social media engagement, thereby exhibiting lower levels of social media addiction.(35)

Finally, Cronbach’s alpha and McDonald’s Omega for the TTAS-SF was 0.805 and 0.817, respectively. Also, Cohen’s kappa for the six items ranged from 0.789 to 0.905 (p < 0.001 in all cases), while intraclass correlation coefficient was 0.993 (p < 0.001). Thus, the reliability of the TTAS-SF was excellent.

## Limitations

This study presents several limitations. First, a convenience sampling method was employed, which restricts the generalizability of the findings. Future research should utilize more representative samples and explore different populations (e.g., students, adolescents) to further validate the TTAS-SF. Nevertheless, the psychometric analysis remains robust, as the sample size met all methodological requirements. Second, the study was not conducted in clinical settings; therefore, caution is advised when applying these findings in diagnostic contexts. Research conducted under well-controlled clinical conditions would provide valuable additional insights. Third, self-report instruments were used to assess the concurrent validity of the TTAS-SF, introducing the possibility of information bias. Finally, concurrent validity was examined through correlations with three scales; future studies should incorporate additional measures to strengthen the validation process.

## Conclusions

To the best of our knowledge, the TTAS is the first instrument specifically designed to measure TikTok addiction among users. Following comprehensive reliability and validity analyses, the short version of the TTAS (i.e., the TTAS-SF) was found to be a very brief, concise, user-friendly tool with strong psychometric properties. Our findings indicate that the TTAS-SF is a one factor 6-item scale, reflecting the core components of addiction: salience, mood modification, tolerance, withdrawal, conflict, and relapse. Consequently, the TTAS-SF represents a timely and practical measure for assessing TikTok addiction and identifying high-risk individuals in both community and educational settings. In light of the study’s limitations, we recommend translating and validating the TTAS-SF across different languages and populations to further establish its reliability and validity. The TTAS-SF has the potential to serve as an effective instrument for detecting TikTok addiction and may assist policymakers, health educators, clinicians, and researchers in identifying vulnerable groups.

## Data Availability

All data produced are available online at https://doi.org/10.6084/m9.figshare.26314522.v1

https://doi.org/10.6084/m9.figshare.26314522.v1

## References

1. Montag C, Yang H, Elhai JD. On the Psychology of TikTok Use: A First Glimpse From Empirical Findings. Frontiers in Public Health. 2021;9: 641673. 10.3389/fpubh.2021.641673.

2. Arrivillaga C, Rey L, Extremera N. A mediated path from emotional intelligence to problematic social media use in adolescents: The serial mediation of perceived stress and depressive symptoms. Addictive Behaviors. 2022;124: 107095. 10.1016/j.addbeh.2021.107095.

3. Bányai F, Zsila Á, Király O, Maraz A, Elekes Z, Griffiths MD, et al. Problematic Social Media Use: Results from a Large-Scale Nationally Representative Adolescent Sample. Jiménez-Murcia S (ed.) PLOS ONE. 2017;12(1): e0169839. 10.1371/journal.pone.0169839.

4. Sindermann C, Elhai JD, Montag C. Predicting tendencies towards the disordered use of Facebook’s social media platforms: On the role of personality, impulsivity, and social anxiety. Psychiatry Research. 2020;285: 112793. 10.1016/j.psychres.2020.112793.

5. Keles B, McCrae N, Grealish A. A systematic review: the influence of social media on depression, anxiety and psychological distress in adolescents. International Journal of Adolescence and Youth. 2020;25(1): 79–93. 10.1080/02673843.2019.1590851.

6. Kuss D, Griffiths M, Karila L, Billieux J. Internet Addiction: A Systematic Review of Epidemiological Research for the Last Decade. Current Pharmaceutical Design. 2014;20(25): 4026–4052. 10.2174/13816128113199990617.

7. Xanidis N, Brignell CM. The association between the use of social network sites, sleep quality and cognitive function during the day. Computers in Human Behavior. 2016;55: 121–126. 10.1016/j.chb.2015.09.004.

8. Smith T, Short A. Needs affordance as a key factor in likelihood of problematic social media use: Validation, latent Profile analysis and comparison of TikTok and Facebook problematic use measures. Addictive Behaviors. 2022;129: 107259. 10.1016/j.addbeh.2022.107259.

9. Casale S, Rugai L, Fioravanti G. Exploring the role of positive metacognitions in explaining the association between the fear of missing out and social media addiction. Addictive Behaviors. 2018;85: 83–87. 10.1016/j.addbeh.2018.05.020.

10. Tarafdar M, Maier C, Laumer S, Weitzel T. Explaining the link between technostress and technology addiction for social networking sites: A study of distraction as a coping behavior. Information Systems Journal. 2020;30(1): 96–124. 10.1111/isj.12253.

11. Katsiroumpa A, Katsiroumpa Z, Koukia E, Mangoulia P, Gallos P, Moisoglou I, et al. Association Between Problematic TikTok Use and Procrastination, Loneliness, and Self-Esteem: A Moderation Analysis by Sex and Generation. European Journal of Investigation in Health, Psychology and Education. 2025;15(10): 209. 10.3390/ejihpe15100209.

12. Bilali A, Katsiroumpa A, Koutelekos I, Dafogianni C, Gallos P, Moisoglou I, et al. Association Between TikTok Use and Anxiety, Depression, and Sleepiness Among Adolescents: A Cross-Sectional Study in Greece. Pediatric Reports. 2025;17(2): 34. 10.3390/pediatric17020034.

13. Katsiroumpa A, Moisoglou I, Gallos P, Katsiroumpa Z, Konstantakopoulou O, Tsiachri M, et al. Problematic TikTok Use and Its Association with Poor Sleep: A Cross-Sectional Study Among Greek Young Adults. Psychiatry International. 2025;6(1): 25. 10.3390/psychiatryint6010025.

14. Galanis P, Katsiroumpa A, Katsiroumpa Z, Mangoulia P, Gallos P, Moisoglou I, et al. Association between problematic TikTok use and mental health: A systematic review and meta-analysis. AIMS Public Health. 2025;12(2): 491–519. 10.3934/publichealth.2025027.

15. Varona MN, Muela A, Machimbarrena JM. Problematic use or addiction? A scoping review on conceptual and operational definitions of negative social networking sites use in adolescents. Addictive Behaviors. 2022;134: 107400. 10.1016/j.addbeh.2022.107400.

16. Andreassen CS, Torsheim T, Brunborg GS, Pallesen S. Development of a Facebook Addiction Scale. Psychological Reports. 2012;110(2): 501–517. 10.2466/02.09.18.PR0.110.2.501-517.

17. Galanis P, Katsiroumpa A, Moisoglou I, Konstantakopoulou O. The TikTok Addiction Scale: Development and validation. AIMS Public Health. 2024;11(4): 1172–1197. 10.3934/publichealth.2024061.

18. World Medical Association. World Medical Association Declaration of Helsinki: Ethical Principles for Medical Research Involving Human Subjects. JAMA. 2013;310(20): 2191. 10.1001/jama.2013.281053.

19. Yusoff MSB, Arifin WN, Hadie SNH. ABC of Questionnaire Development and Validation for Survey Research. Education in Medicine Journal. 2021;13(1): 97–108. 10.21315/eimj2021.13.1.10.

20. Brown T. Confirmatory Factor Analysis for Applied Research.. 2nd edn New York: The Guilford Press; 2015.

21. Hu L, Bentler PM. Fit indices in covariance structure modeling: Sensitivity to underparameterized model misspecification. Psychological Methods. 1998;3(4): 424–453. 10.1037/1082-989X.3.4.424.

22. Baumgartner H, Homburg C. Applications of structural equation modeling in marketing and consumer research: A review. International Journal of Research in Marketing. 1996;13(2): 139–161. 10.1016/0167-8116(95)00038-0.

23. Andreassen CS, Billieux J, Griffiths MD, Kuss DJ, Demetrovics Z, Mazzoni E, et al. The relationship between addictive use of social media and video games and symptoms of psychiatric disorders: A large-scale cross-sectional study. Psychology of Addictive Behaviors. 2016;30(2): 252–262. 10.1037/adb0000160.

24. Kroenke K, Spitzer RL, Williams JBW, Lowe B. An Ultra-Brief Screening Scale for Anxiety and Depression: The PHQ-4. Psychosomatics. 2009;50(6): 613–621. 10.1176/appi.psy.50.6.613.

25. Rammstedt B, John OP. Measuring personality in one minute or less: A 10-item short version of the Big Five Inventory in English and German. Journal of Research in Personality. 2007;41(1): 203–212. 10.1016/j.jrp.2006.02.001.

26. Katsiroumpa A, Katsiroumpa Z, Koukia E, Mangoulia P, Gallos P, Moisoglou I, et al. Bergen Social Media Addiction Scale: Translation and validation in Greek. International Journal of Caring Sciences. 2025;18(2): 661–671. https://osf.io/93rns_v1

27. Karekla M, Pilipenko N, Feldman J. Patient Health Questionnaire: Greek language validation and subscale factor structure. Comprehensive Psychiatry. 2012;53(8): 1217–1226. 10.1016/j.comppsych.2012.05.008.

28. Soto CJ, John OP. The next Big Five Inventory (BFI-2): Developing and assessing a hierarchical model with 15 facets to enhance bandwidth, fidelity, and predictive power. Journal of Personality and Social Psychology. 2017;113(1): 117–143. 10.1037/pspp0000096.

29. Bland JM, Altman DG. Statistics notes: Cronbach’s alpha. BMJ. 1997;314(7080): 572–572. 10.1136/bmj.314.7080.572.

30. De Vaus D. Surveys in social research.. 5th edn London: Routledge; 2004.

31. Cunningham S, Hudson CC, Harkness K. Social Media and Depression Symptoms: a Meta-Analysis. Research on Child and Adolescent Psychopathology. 2021;49(2): 241–253. 10.1007/s10802-020-00715-7.

32. Hussain Z, Wegmann E, Yang H, Montag C. Social Networks Use Disorder and Associations With Depression and Anxiety Symptoms: A Systematic Review of Recent Research in China. Frontiers in Psychology. 2020;11: 211. 10.3389/fpsyg.2020.00211.

33. Correa T, Hinsley AW, D. Zúñiga HG. Who interacts on the Web?: The intersection of users’ personality and social media use. Computers in Human Behavior. 2010;26(2): 247–253. 10.1016/j.chb.2009.09.003.

34. Kuss DJ, Griffiths MD. Online Social Networking and Addiction—A Review of the Psychological Literature. International Journal of Environmental Research and Public Health. 2011;8(9): 3528–3552. 10.3390/ijerph8093528.

35. Wilson K, Fornasier S, White KM. Psychological predictors of young adults’ use of social networking sites. Cyberpsychology, Behavior and Social Networking. 2010;13(2): 173–177. 10.1089/cyber.2009.0094.

